# Rapid, Near Point-of-Care Assay for HLA-B*57:01 Genotype Associated with Severe Hypersensitivity Reaction to Abacavir

**DOI:** 10.1101/2021.05.26.21257187

**Authors:** Jackson J. Wallner, Ingrid A. Beck, Nuttada Panpradist, Parker S. Ruth, Humberto Valenzuela-Ponce, Maribel Soto-Nava, Santiago Ávila-Ríos, Barry R. Lutz, Lisa M. Frenkel

## Abstract

**Objective:** The nucleoside reverse transcriptase inhibitor abacavir is commonly used to treat young children with HIV infection. Abacavir can trigger a severe hypersensitivity reaction in people who are homozygous or heterozygous for *HLA-B*57:01*. Testing for *HLA-B*57:01* prior to abacavir initiation is standard-of-care in high-resource settings, but current tests are too costly for resource-limited settings. To address this gap, we developed an inexpensive, simple-to-use rapid assay to detect *HLA-B*57:01*.

**Methods:** We designed and optimized a multiplexed PCR to amplify *HLA-B*57* subtypes and the human beta-globin gene. Subsequently, probes annealed to the amplicon and were ligated when specific for the *HLA-B*57:01* allele. Ligated products were detected by immunocapture in a lateral flow strip. Cell lines with known HLA genotypes were used to optimize the assay. The assay was then evaluated by comparing the genotype of clinical specimens (n=60) enriched for individuals with *HLA-B*57:01* by the new assay to that from sequencing.

**Results:** The optimized multiplex PCR for *B*57* and beta-globin resulted in a 40-minute, 35-cycle amplification, followed by a 20-minute ligation reaction and 15-minute detection step. Evaluation of the *HLA-B*57:01* oligonucleotide ligation assay using clinical specimens had a sensitivity of 100% (n=27/27 typed as *B*57:01*) and specificity of 100% (n=33/33 typed as non-*B*57:01*) by visual interpretation of lateral flow strips.

**Conclusions:** This rapid and economical assay can accurately detect the presence of *HLA-B*57:01* in clinical specimens. Use of this assay could expand access to *HLA-B*57:01* genotyping and facilitate safe same-day initiation of abacavir-based treatment.

## Introduction

Abacavir (**ABC**) is a nucleoside reverse transcriptase inhibitor used in combination therapy to treat HIV [1–3]. Despite high tolerability in most patients, ABC triggers a hypersensitivity reaction in 5-8% of people within 6 weeks of treatment initiation, characterized by fever, rash, nausea, and respiratory distress. Treatment continuation or drug re-initiation can be fatal [4]. The risk of developing a hypersensitivity reaction to ABC is strongly associated with the *HLA*-*B*57:01* allele [5]. The allele frequency of *HLA*-*B*57:01* varies by ethnicity; ranging from 0-3% in African and South American populations, 6-10% in Caucasian populations and up to 19% in Southwest Asian populations [6]. Screening for *HLA*-*B*57:01* has become standard-of-care prior to ABC use in high-resource communities [3].

Current tests for ABC hypersensitivity detect the *HLA-B*57:01* subtype by allele-specific PCR or sequencing. Allele-specific PCR uses primers [7, 8] to specifically amplify the *HLA-B*57:01* allele, requiring a standard thermocycler and electrophoresis equipment [7] (cost estimated at US$ 1,500), or real-time fluorescence instruments [9] (cost estimated at US$ 15,000-50,000). Sanger sequencing or next generation sequencing [7, 10] require even more expensive equipment (>US$ 100,000). Due to high-cost instruments, most *HLA-B*57:01* testing is performed at centralized laboratories, which requires shipping of specimens. Slow turn-around-times of 3 days to weeks can delay treatment initiation. Tests that could be performed inexpensively without the need for sequencing technology could increase the safe use of ABC in settings where screening for *HLA-B*57:01* is otherwise cost-prohibitive.

Here we report the development and validation of a lateral flow-based oligonucleotide ligation assay (**OLA**) that detects a polymorphism differentiating *HLA*-*B*57:01* from other *B*57* subtypes. *B*57* subtypes are amplified from genomic DNA followed by annealing to two probes that bisect the *HLA-B*57:01* allele. A ligase joins the two probes only when the probe sequences match the amplified product at the ligation site. The ligated product is detected by immunocapture on a single-use lateral flow strip. This assay only requires a standard thermocycler and microcentrifuge (cost estimated <US$ 1,000), which are relatively inexpensive and common in small labs, allowing use in near point-of-care settings.

## Materials and Methods

### Study design and population

PCR primers were chosen to amplify the region of chromosomal DNA encoding *HLA-B*57* subtypes [7]. OLA probes were designed to detect *HLA-B*57:01* but not other *HLA-B*57* subtypes. The PCR for *HLA-B*57* was multiplexed with primers for human beta-globin as a positive control for DNA extraction, amplification and ligation. Assay optimization used DNA from five cell lines (LADA, MYE 2004, FH18, 32/32, DBB; from the Fred Hutchinson International Histocompatibility Working Group, Seattle, WA) including *HLA-B*57:01, B*57:02, B*57:03* and an *B*57* negative control. Clinical validation used 60 banked specimens, including 52 Mexican [10] and 8 Seattle Primary Infection Cohort [11] DNA specimens. The Mexican specimens were enriched for *HLA-B*57:01* and other common *B*57* subtypes (i.e., *B*57:02/03*) compared to the Mexican population and were previously typed using next generation sequencing (Trusight HLA v2 Sequencing Panel on MiniSeq platform [Illumina, San Diego, CA], and Assign v2.0 software was used to assign HLA subtypes) at the Centre for Research in Infectious Diseases (CIENI) of the National Institute of Respiratory Diseases (INER) in Mexico City. Studies collecting these samples and those from the Seattle Primary Infection Cohort were approved by respective Ethics Committees. De-identified DNA from Mexican specimens were sent to Seattle for testing. Following blinded testing by the *HLA-B*57:01* OLA assay, the results were compared to the genotypes determined by sequencing.

### Sample preparation

DNA was extracted from buffy coat (200μL) or PBMCs (peripheral blood mononuclear cells, ∼6 million cells) using the QIAmp DNA Blood Mini Kit (Qiagen, Valencia; CA) according to manufacturer’s specifications.

### Primers and PCR

The primer sets coamplify a fragment comprising 161 nucleotides of the human beta-globin gene (forward primer: 5’-GGGATCTGTCCACTCCTGATGCTGT; reverse primer: 5’-ATCCACGTGCAGCTTGTCACAGTG) and a region (179bp) specific to *B*57* subtypes using a modification of primers previously published [7] (forward primer: 5’-CCAGGGTCTCACATCATCCAGGT; reverse primer: 5’-CGCCTCCCACTTGCGCTGGG) (Integrated DNA Technology, Coralville, Iowa). The PCR was performed in a 25 µL reaction containing 0.625 U Terra PCR Direct Polymerase (Takara Bio Inc., Shiga, Japan), 0.4 µM of each primer, 12.5 µL Terra buffer, and 2.5 µL of extracted DNA (5-10 ng total DNA). Two-step PCR conditions are as follows: 30 seconds at 95°C, 35 cycles with denaturation for 15 seconds at 95°C followed by annealing for 30 seconds at 68°C, with a final extension at 68°C for 5 minutes.

### Oligonucleotide ligation assay (OLA)

Oligonucleotide probes for *HLA-B*57:01* target a cytosine nucleotide at position 1094 of the *B*57:01* allele (reference sequence *HLA-B*57:01:01*, GenBank Accession number AJ458991), but not other *B*57* types [7]: *B*57*-probe-1: 5’-FAM-TCCTCC GCGGGCATGACCAGTC; *B*57*-probe-2: 5’-Phosphorylation-YGCCTACGACGGCAAGGATTACA–Biotin. Beta-globin probes ligate when annealed to the beta-globin amplicon and serve as an amplification and ligation control: beta-globin-probe-1: 5’-Digoxigenin-CTCGGTGCCTTTAGTGATGGC CTG; beta-globin-probe-2: 5’-Phosphorylation-GCTCACCTGGACAACCTCAAGGG–Biotin. Two µL of amplicon from the multiplex PCR are added directly into 20 µL of ligation mix containing 1.33 U Ampligase DNA Ligase (Lucigen, Middleton, WI), 12.5mM KCl, 1mM NAD, 1x ligase buffer (20mM Tris-HCl, 10mM MgCl_2_, 1mM DDT), 0.08% Triton(tm)X-100, 16.7nM *B*57* probes, 4.2nM beta-globin-probe-1 and 16.7nM beta-globin-probe-2. The ligation reaction conditions are: 10 cycles with denaturation at 94°C for 30 seconds followed by ligation for 30 seconds at 50°C.

### Detection of ligation products

Plate-based OLA: Ligated products are captured on a streptavidin-coated 96-well plate (Sigma-Aldrich, St Louis, USA) for 1 h at room temperature and then denatured by washing with dilute NaOH (0.01 N NaOH and 0.05% Tween®20), followed by incubation with anti-fluorescein and anti-digoxigenin antibodies (Sigma, St. Louis, MO). Secondary reporters are added sequentially for *B*57:01* and beta-globin, and the optical density (**OD**) was measured using a Spectramax i3x plate reader (Molecular Devices, San Jose, CA) at 405 and 450 nm, respectively [12]. Samples with high signal (OD > 1.0) at both wavelengths were considered *B*57:01* positive. Samples with low (OD < 0.15) signal at 405nm and high signal at 450 nm were considered negative.

Paper-based OLA: Unlabeled oligonucleotides with identical sequences to the ligation probes were added at 40-fold higher concentration and denatured at 90°C for 30 seconds. The 26.8 µL ligation reaction is then added onto lateral flow detection strips and after 5 minutes is chased by 43 µL buffer containing anti-biotin antibodies conjugated gold nanoparticles (OD=0.7 at 520 nm). Ten minutes after the addition of the gold buffer, the strips were visually inspected for appearance of precipitated bands to determine genotype or by scanner and in-house software.

### Preparation of lateral flow test

Anti-fluorescein (Southern Biotech, Birmingham, AL) and anti-digoxigenin (Novus Biologicals, Centennial CO, USA) antibodies to capture ligated products complementary to *HLA-B*57:01* and beta-globin, respectively, were immobilized on lateral flow strips, and BSA-conjugated biotin (Sigma Aldrich, Saint Louis, MO) was used as a flow control as previously described [13–15].

## Results

### Development of the HLA-B*57:01 OLA assay

We developed an assay that co-amplifies the beta-globin gene and the *B*57* allele from 100% of specimens previously typed as *B*57* positive using supplies costing <US$ 6/specimen. *B*57:01* probe concentrations and ligation conditions were optimized to maximize signal to noise ratio in a 96-well ELISA-based OLA using cell lines with known *B*57:01/02/03* subtypes or no *B*57* variants. The OLA was then adapted to a lateral flow strip format with visual detection. In addition, in-house software was used to quantify band signal intensities from scanned images (Fig. 1).

**Fig. 1.**
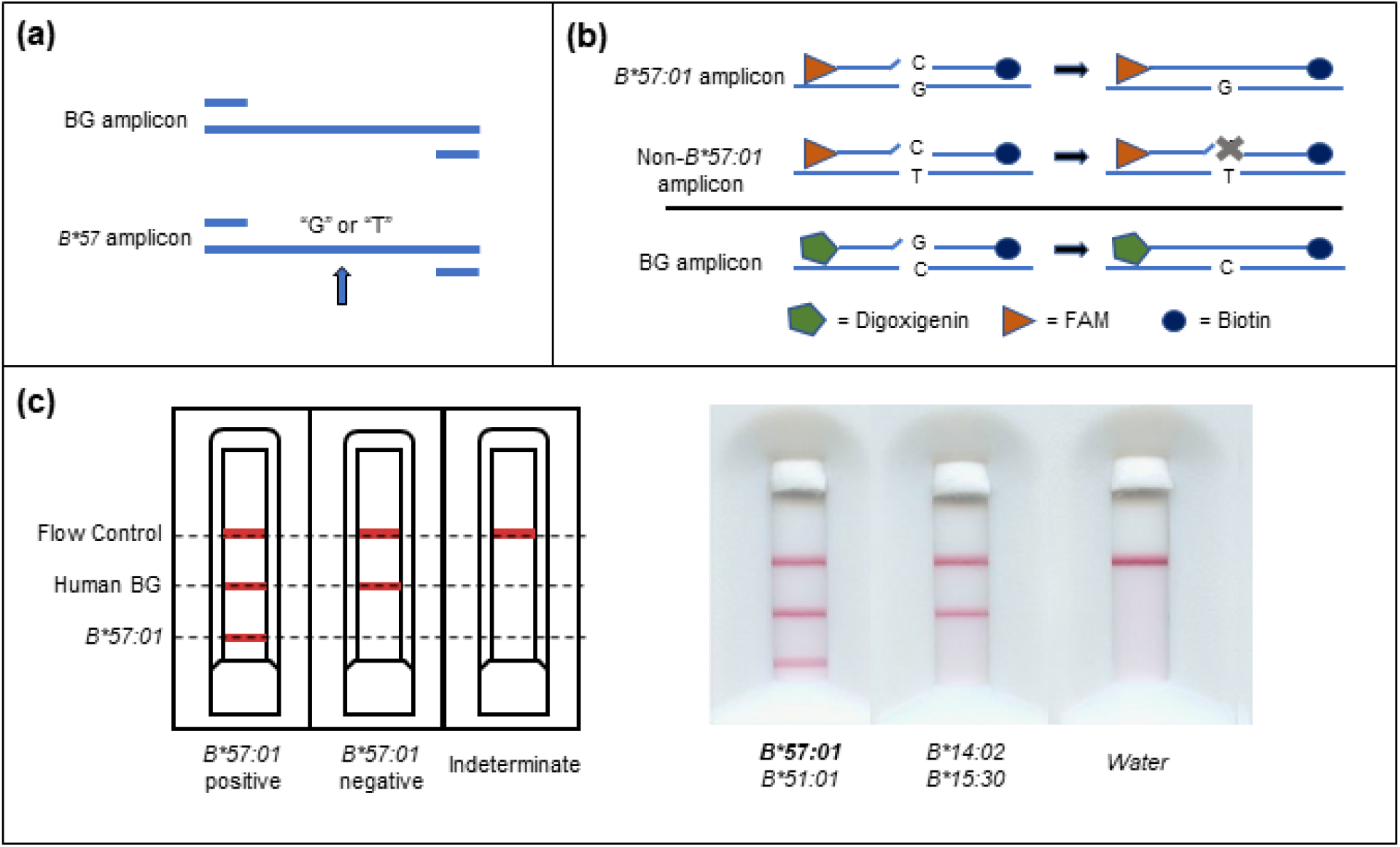
Oligonucleotide ligation assay for detection of *HLA-B*57:01*. (a) Amplification: Extracted DNA is added to the PCR reaction mixture for multiplexed amplification of human beta-globin (**BG**) and *B*57* gene fragments. (b) Ligation: Labeled *B*57:01* probes are selectively ligated together in the presence of amplicons with the *B*57:01* allele. BG probes ligate in the presence of the amplified BG region of the somatic genome and serve as a (+) control. (c) Lateral flow detection: Chip layout and scanned images of participant sample assays. Appearance of a red line at “Flow Control” indicates adequate flow of reagents on the single-use cartridge; a red line at “Human BG” indicates presence of human BG amplicon, and confirms adequate DNA extraction, PCR amplification and ligation; a red line at “*B*57:01*” indicates detection of *B*57:01*, either a specimen that is homozygous or heterozygous for the *B*57:01* subtype. HLA-B* types determined by Illumina sequencing are shown for two participants’ samples tested by OLA.

### Validation of OLA for detection of HLA-B*57:01 subtypes using clinical specimens

DNA extracted from 60 clinical samples were tested for *B*57:01* by the lateral flow-based assay and classified as positive or negative for both beta globin and *B*57:01* bands by visual assessment. Compared to HLA typing by sequencing, visual inspection of the OLA classified as positive all 27 specimens heterozygous for *B*57:01* (100% sensitivity [95% confidence interval: 87.2-100%]) (Table 1). The remaining 33 specimens were correctly classified as *B*57:01* negative (100% specificity [95% confidence interval: 89.4-100%]). All samples were classified as positive for the human beta globin control.

**Table 1.** HLA-*B*57:01* typing results by OLA and Illumina sequencing in a Mexican population enriched for HLA-*B*57* and Seattle Primary Infection Cohort

| **HLA-B Typing Results** |  |  |  |  |
| --- | --- | --- | --- | --- |
| HLA-B type by Illumina Sequencing | Number of specimens | Expected outcome by OLA | Correct OLA outcome by visual interpretation | Correct OLA outcome by in-house software |
| B*57:01 | 27 | + | 27 | 27 |
| B*57:02 | 1 | - | 1 | 1 |
| B*57:03 | 9 | - | 9 | 9 |
| Non-B*57<sup>a</sup> | 23 | - | 23 | 23 |

<sup>a</sup>Non-*B\*57* types tested included - *B\*07, 08, 13, 14, 15, 18, 35, 38, 39, 40, 41, 42, 44, 45, 48, 49, 51, 52*.

### Validation of OLA by In-House Software

To avoid human error, the lateral flow strips can be analyzed using in-house Python software, based on an algorithm previously described [14, 15]. The software automatically detected bands in the paper strips and quantified signal intensity by subtracting brightness within band and adjacent background regions. By the in-house software, all 60 specimens were correctly classified as *B*57:01* positive or negative (100% sensitivity and specificity).

## Discussion

We developed and evaluated an inexpensive and relatively rapid assay that accurately detects the *HLA-B*57:01* subtype, associated with a hypersensitivity reaction to ABC, and discriminates it from similar common *B*57* subtypes (i.e., *B*57:02* and *B*57:03*). Assay operation requires minimal supplies and relatively inexpensive equipment and delivers results in less than 2 hours (i.e., 30-minute DNA extraction, 40-minute PCR, 20-minute ligation, and 15-minute detection). Results are simple to analyze by the unaided eye or using a scanner and our in-house software [14, 15].

This assay showed high sensitivity and specificity for detection of *HLA*-*B*57:01* in clinical samples compared to HLA typing by sequencing. Performance of the OLA was similar to other non-sequencing methods such as allele-specific PCR [7, 9, 16].

This study was limited by a relatively low number of clinical samples tested; however, it was enriched to include all with the *B*57:01* subtype from a HIV cohort of ∼1,800 individuals in Mexico City. A sample panel from more diverse populations would allow a more comprehensive validation of primers and probes and discrimination of *B*57:01* from other HLA subtypes. The primers and probes used in this assay are complementary at the site of ligation to sequences described for two other alleles (*B*57:08* and *B*55:14*) [7], which would likely result in a false positive result for a non-*B*57:01* specimen. However, the frequencies of these two HLA subtypes across most of the world’s populations are much lower than that of *HLA*-*B*57:01* [17, 18, 19].

In this study we only tested DNA extracted from blood samples as we did not have access to other sample types. The use of an inhibitor-resistant polymerase for PCR may facilitate direct input from fingerstick blood or buccal swabs, thus eliminating the need for DNA extraction or venipuncture, respectively [20]. A change in specimen type and use of direct-to-PCR samples would reduce reagent costs to US$ 2.05/specimen. To increase ease-of-use, test reagents can be lyophilized for single use aliquots [15, 21, 22]. These improvements would simplify the procedure, shorten processing time and aid implementation in near point-of-care settings.

## Conclusion

ABC is recommended as first-line HIV treatment in children by multiple experts [3, 23]. However its safe use is limited by the high cost and limited access to genotypic assays to detect *B*57:01*, which increases the risk of a severe hypersensitivity reactions [4, 5]. The simple, rapid and inexpensive *B*57:01* OLA described here allow testing for this HLA subtype in low-resource settings, which could facilitate safer same-day initiation of ABC-based treatment.

## Data Availability

Genbank submission pending

## Acknowledgments

Support for this project came from the National Institutes of Health (NIH) including R01 AI110375 (LMF); R01 AI145486 (BRL); the Clinical and Retrovirology Research Core and the Molecular Profiling and Computational Biology Core of the University of Washington Fred Hutch Center for AIDS Research [P30 AI027757]; and the International Maternal Pediatric Adolescent AIDS Clinical Trials Network (IMPAACT). IMPAACT is funded by the National Institute of Allergy and Infectious Diseases (NIAID) with co-funding from the Eunice Kennedy Shriver National

Institute of Child Health and Human Development (NICHD) and the National Institute of Mental Health (NIMH), all components of NIH under Award Numbers UM1AI068632 (IMPAACT LOC), UM1AI068616 (IMPAACT SDMC) and UM1AI106716 (IMPAACT LC), and by NICHD contract number HHSN275201800001I. The work was supported in part by funds from the Canadian Institutes of Health Research (grants PJT-148621 and PJT-159625), received by SAR, as well as funds from the Mexican Government (Comisión de Equidad y Género de las Legislaturas LX-LXI y Comisión de Igualdad de Género de la Legislatura LXII de la H. Cámara de Diputados de la República Mexicana), received by SAR. The content is solely the responsibility of the authors and does not necessarily represent the official views of the funders.

## Author Contributions

J.J.W. developed and optimized the assay and performed blinded sample testing. I.A.B. and N.P. developed the assay platform and provided writing and project guidance. N. P. and P.S.R. manufactured the tests and performed software analysis for the project. H.V-P., M.S-N., S.A-R. collected and provided samples for blinded testing and sequencing data. B.R.L., S.A-R and L.M.F. oversaw the project and provided guidance and feedback.

